# Mitigation Strategies and Compliance in the Covid-19 Fight; How Much Compliance is Enough?

**DOI:** 10.1101/2020.09.07.20189449

**Authors:** Swati Mukerjee, Clifton Chow, Mingfei Li

## Abstract

Today, with only 4% of the world’s population, the U.S. is bearing a disproportionate share of COVID-19 infections. Seeking to understand this puzzle, we investigate how mitigation strategies and compliance can work together (or in opposition) to reduce (or increase) the spread of COVID-19 infection. Drilling down to the state level, we create specific state indices suitable for the U.S. to measure the degree of strictness of public mitigation measures. In this, we build on the Oxford Stringency Index. A modified time-varying SEIRD model, incorporating this Stringency Index as well as a Compliance Indicator to reduce the transmission, is then estimated with daily data for a sample of 6 U.S. states. These are New York, New Hampshire, New Mexico, Colorado, Texas, and Arizona. We provide a simple visual policy tool to evaluate the various combinations of mitigation policies and compliance that can reduce the basic reproduction number to less than one; this is the acknowledged threshold in the epidemiological literature to control the pandemic. States successful in combating the pandemic were able to achieve a suitable combination. Understanding of this relationship by the public and policy makers is key to controlling the pandemic. This tool has the potential to be used in a real-time, dynamic fashion for flexible policy options.

## 1. INTRODUCTION

The COVID-19 pandemic has resulted in 20 million infections and 760,213 deaths globally (Johns Hopkins University & Medicine, 2020). Of these, the U.S. has contributed over 5 million cases, with 167,253 deaths. This has happened while testing has accelerated in the U.S. According to the WHO (World Health Organization), the positivity rate in testing (the percentage of tests conducted that are positive for COVID-19) should be between 3% and 5%. However, before reopening is considered, the positivity rate should be 5% or below for at least 14 days (WHO, 2020). On August 13, 2020, the U.S. had an overall positive rate of 7.5% (Ritchie, et al., 2020) which is well above the upper bound recommended. Why is the U.S. then bearing such a disproportionate burden of infective cases when it has only 4.25% of the world’s population? (U.S. Census Bureau, 2020)

Clues as to why the overall infections are so high can only be seen by disaggregating to the state level where we encounter considerable heterogeneity. On August 16, only 17 states had met the positivity recommendations (Johns Hopkins University & Medicine, 2020). Further disparities emerge from the selective adoption by the states (to varying degrees of strictness) of the community mitigation strategies recommended by the CDC in the absence of a proven and widely available therapy or vaccine (Centers for Disease Control and Prevention, 2020). Adding to this confusing mosaic are the diverse degrees of compliance by the public in each state.

The objective of this study is to investigate how mitigation strategies and compliance can work together (or in opposition) to reduce (or increase) the spread of infection. To accomplish this, we build an epidemiological model that specifically takes into account not only the community mitigation strategies that slow the spread of the virus, but also compliance^1^ by the public. This model is applied to a sample of 6 U.S. states (New York, New Hampshire, New Mexico, Colorado, Texas, and Arizona) chosen for three reasons. They have had varying success in flattening the infection curve; they have the daily data on recoveries that is needed for estimation; they have the advantage of having a range of positivity ratios^2^. In view of our estimation results, we offer some practical policy tools and recommendations to aid public policy.

As of today, while the race for a vaccine proceeds, mitigation actions are the primary bulwark against COVID-19^3^. These measures can move from the individual level like washing hands, wearing face masks^4^, and maintaining physical distance to those imposed by authorities such as restrictions on gatherings, school and workplace closings etc. The Oxford University Blavatnik School of Government maintains a website that has created an index to capture such strategies (Hale, et al., 2020). With data from more than 160 countries including the U.S., it has calculated a Government Response Stringency Index (GRSI) for each country using 9 indicators of mitigation such as school closings, restrictions on gatherings and so on (Hale et. al. 2020) See Appendix A for the individual indicators. This GRSI is scaled from 0 to 100. As of July 31, 2020, the Index for the U.S. is 69.0 and to give this some context, for Canada it is 67.13, Australia is 68.06, China is 81.94. Very recently, it has also constructed Stringency Indices for each U.S. state (Hale, et al., 2020B).

The national GRSI has been used by researchers (Jaytilleke et. al. 2020) and has also been used in a modified version applied to Brazil (Barberia et. 2020). We are building on the methodology proposed by the Oxford team for the state indices they have created. On examining the individual elements that go into the state indices calculated by the Oxford index, we realized that certain changes were needed to construct state indices tailored to U.S. state conditions. By modifying individual elements we created another set of state-specific Stringency Indices that we call the Bentley State Stringency Index (BSI). We have therefore built on and extended the contribution by the Oxford University Martin School.

As the purpose of mitigation measures is to slow down the spread of infection, we introduced an exponential Mitigation Function on the transmission term in a time-varying SEIRD model. This Mitigation Function, incorporating the Bentley State Stringency Index (BSI), plays a crucial role in slowing down the progression of the disease, provided there is compliance. The latter is represented by an estimable Compliance Indicator (CI) that captures the average degree of compliance in each state. The Compliance Indicator thus modifies the effect of the BSI. It can allow the BSI to work at its full potential in reducing infection or it can progressively choke off completely the effect of mitigation policies as compliance by the public moves to zero. Sheikh et. al. 2020 outlined some indirect ways in which one may assess the degree of compliance such as by cell phone GPS data or traffic congestion and public transport usage. Our approach provides a data-based estimation of the degree of compliance that is state specific. To the best of our knowledge, our approach of employing a Mitigation Function with a tailored, state specific Stringency Index and a Compliance Indicator has not been done before.

The estimated Compliance Indicator was then used with varying values of the Stringency Index to see how the basic reproductive number R_0_^5^ could be brought to a level less than 1^6^ in each of the states studied. This provided some answers as to why some states have not been successful in controlling infections. Our work focuses on the importance of the Mitigation Function with its two important components: the BSI and the Compliance Indicator. We argue that this, too, can be a useful metric to be watched by policy makers. We have given the method by which it can be calculated and shown the value of tracking it to ensure that it is within acceptable limits. This metric can indicate the minimum level of compliance needed to control the epidemic given a particular level of stringency. The visual tool that we present also has the advantage of being easy for the public to understand.

## 2. DATA, VARIABLES AND DESCRIPTIVE STATISTICS

Our work is based on multiple data sources. The New York Times repository of coronavirus data on GitHub and state level data from the various state web portals^7^ were used. Though these data sets begin from Jan 1 2020, COVID-19 infections were not apparent during the early period. March 2 is the earliest date when any observation is available. Beginning from that date, our data runs through July 31 2020. Additional data on average household size^8^ and state population were gathered from the U.S. Department of the Census projections (U.S. Census Bureau, 2020).

We calculated daily cumulative infection (*infection cases)* from the daily numbers of new confirmed cases. Similarly, daily cumulative recovery (*recovery)* and daily cumulative death (*death)* were computed from daily numbers of recoveries and confirmed deaths. All three variables are from the New York Times repository for each state. States may differ on how recovery is defined. For example, Texas calculated recoveries from those who are hospitalized by estimating the proportion of those who are hospitalized for no more than 32 days. To this number they add those who have not been hospitalized but have been infected with Covid-19 for 14 days (State of Texas, 2020). Colorado bases recovery data on those discharged from Covid-19-related hospitalization (State of Colorado, 2020). For New Hampshire, recoveries are estimated from the resolution of Covid-19 fever without the use of fever-reducing medications and improvements in respiratory symptoms (State of New Hampshire, 2020). No definitions have been released by New York, Arizona or New Mexico at the time the manuscript was completed. Due to the uneven quality in the data, there are a few missing daily entries that occur on different days in each state data across the three different variables that we used: *daily new confirmed cases, daily recovery* and *daily deaths*. Together, they comprise 4% of our 2736 data points. These missing entries were imputed by taking the average of the previous seven days as it takes the CDC coders that length of time to record COVID-19 deaths (CDC, 2020); the CDC uses a 7-day moving average to report new daily cases (Stokes, et al., 2020).

### Overview of the six states

According to WHO, to ensure that the testing rate is sufficient, the positivity rate should be between 3 to 5%. As of August 13^th^, only NY, NH, NM and CO are within this threshold. Back in mid-August, AZ had the highest positivity rate of all 50 states (only Puerto Rico has the highest possible, 100%). The trend of new cases per 100,000 shows a wide disparity in controlling the infection among these states. As seen from table 1, NY which started out with the highest rate of 1154.6 per 100,000 people has now the lowest (111.5). On the other hand, Arizona started with 90 and is now at 1302.4. NM and NH started with very similar rates (144.5 and 167.4 respectively), but by June NM’s rate was almost 2 times of NH’s rate.

**TABLE 1.** Overview of States.

| State | Pos. R | New Cases per 100,000* |  |  |  | State imposed Any Restrictions |  |  |  | Average Household Size | Population |
| --- | --- | --- | --- | --- | --- | --- | --- | --- | --- | --- | --- |
|  |  | April | May | June | July | April | May | June | July |  |  |
| TX | 14.99 | 85.6 | 124.8 | 330 | 900 | Yes | Yes | Yes | Yes | 2.86 | 28,995,881 |
| NM | 3.89 | 144.5 | 210.3 | 222.3 | 418.3 | Yes | Yes | Yes | Yes | 2.64 | 2,096,829 |
| AZ | 24.68 | 87.4 | 168.8 | 814.4 | 1302.4 | Yes | Yes | Yes | Yes | 2.69 | 7,278,717 |
| CO | 7.11 | 217.5 | 196.9 | 111.4 | 237.8 | Yes | Yes | Yes | Yes | 2.56 | 5,758,736 |
| NH | 2.03 | 167.4 | 233.1 | 113.3 | 76.9 | Yes | Yes | Yes | Yes | 2.46 | 1,359,711 |
| NY | 1.08 | 1154.6 | 325.9 | 115.0 | 111.5 | Yes | Yes | Yes | Yes | 2.60 | 19,453,561 |

Source: New York Times Repository of COVID-19 Data.
\*This is obtained by dividing total new cases over the whole month by the state population and then multiplying by 100,000.

In addition, the varying trends of infections and recoveries in each state are shown in Figure 1. In some states like Texas, AZ and NM, the infection rate has accelerated from a previously slower rate of increase. However, in each state there is a clear point of inflection that has occurred in June. On the other hand, the remaining 3 states are entering the phase where the infection is flattening at different rates. This is most noticeable in NY.

**Figure 2:**
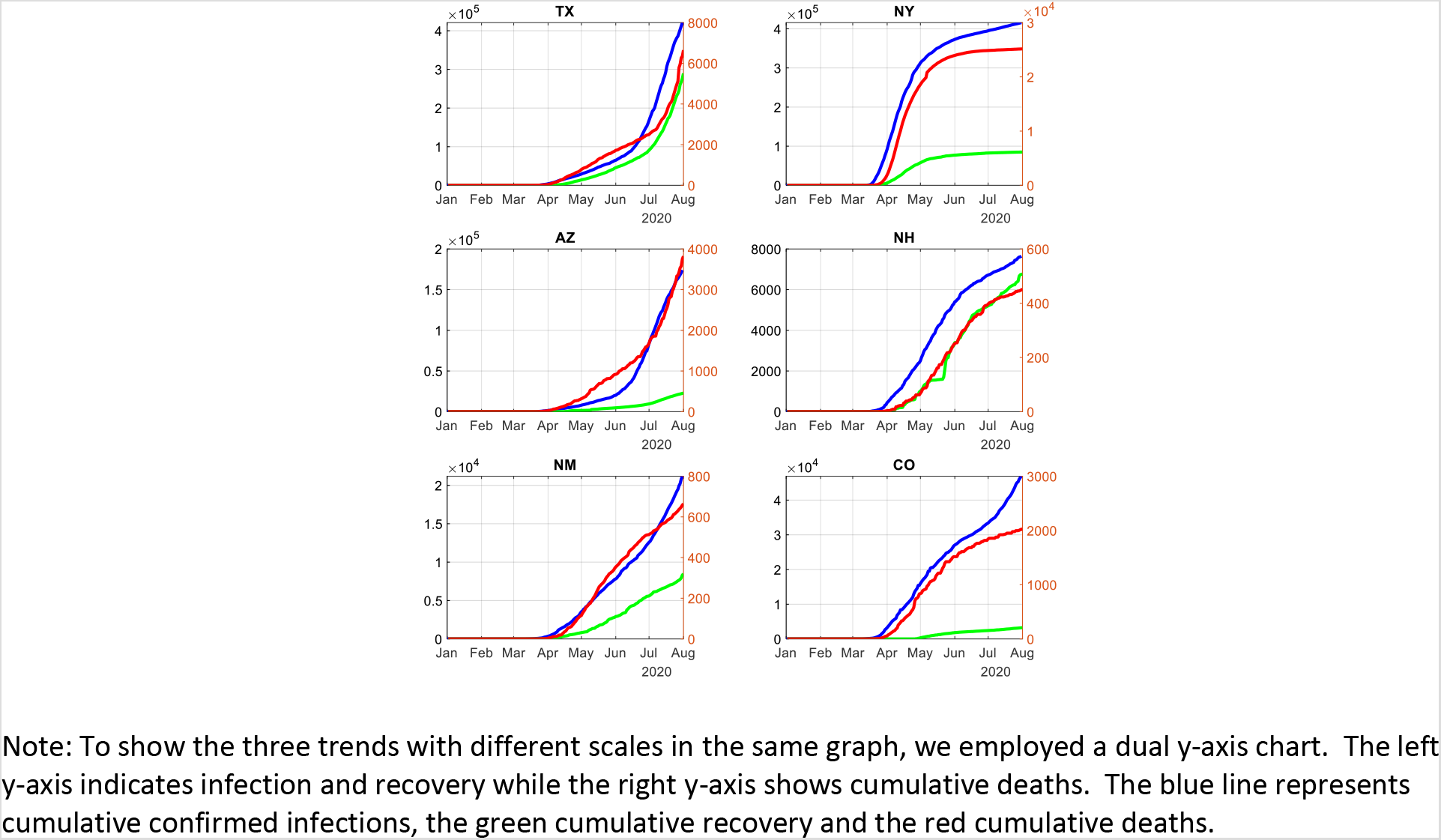
Time series plot for all six states on cumulative confirmed infections, recovery and death. Note: To show the three trends with different scales in the same graph, we employed a dual y-axis chart. The left y-axis indicates infection and recovery while the right y-axis shows cumulative deaths. The blue line represents cumulative confirmed infections, the green cumulative recovery and the red cumulative deaths.

Investigating this disparity is the question that we have set out to explore in this paper.

### The Stringency Index

A Stringency Index was calculated for each state by modifying the Oxford index as shown in Appendix B. Necessary adjustments had to be made in choosing the individual elements of the index. Three items that could not be included by us were restrictions that were not applicable at the State level, or were not applicable to the U.S. in general. These were restrictions on internal travel controls, international transportation, and public officials commenting or coordinating campaigns. However, we needed to add three important restrictions applicable to the U.S. that were not included in the Oxford Index. These were the wearing of face masks^9^, social or physical distancing of 6 feet^10^, nursing home visiting restrictions^11^, and state border restrictions. We adopted the same methodology as given by the Oxford Martin School (Oxford University Martin School, 2020) which we explain in Appendix B.

The BSI was calculated for each state for the entire period. What do these numbers mean? The two extremes of BSI would be 0 and 10. At 0, there are no restrictions. At 10, all the restrictions given in Table 2 (Appendix A) apply at the maximum of the scales applicable to that category. In addition, these restrictions are also applied to the entire state. The actual BSI therefore, will be a combination of the different elements and the scale at which they are applied. For instance, if face masks are recommended but not required, the level of the variable H6 (see Appendix B) becomes 1 instead of the maximum of 2 and the BSI will go down. Therefore, a particular BSI number cannot point to a unique combination of mitigation measures but may be consistent with different combinations of restrictions whether applied to the entire state or to targeted areas.

To further clarify the significance of the BSI, we illustrate it from our calculations using the actual state data from New York and Arizona, two states with very different success in controlling their infection rates. For instance, on the 25 April, the BSI for NY was 6.47 but the next day, 26^th^ April, the BSI jumped to 7.03 because the testing policy was refined to say that anyone showing COVID-19 symptoms should be tested. In Arizona, the BSI on March 30 was 3.75 and the next day it rose to 4.72. The big jump was due to the introduction of new requirements of staying at home and limiting public transportation. On April 30, the BSI was 5.0 and the very next day, May 1, it dropped to 4.4. This is because Arizona removed the restriction and stay at home order ends.

The movement of the BSI for each state is shown in Figure 2. Notice that the 6 states began mitigation interventions around mid-March but subsequently they diverged in terms of timing and extent. By April, CO had the highest BSI, whereas AZ had the lowest number. NY slackened restrictions in June and was slightly below NM, but from mid of April to May, New York had been more restrictive than NM.

**Figure 2.**
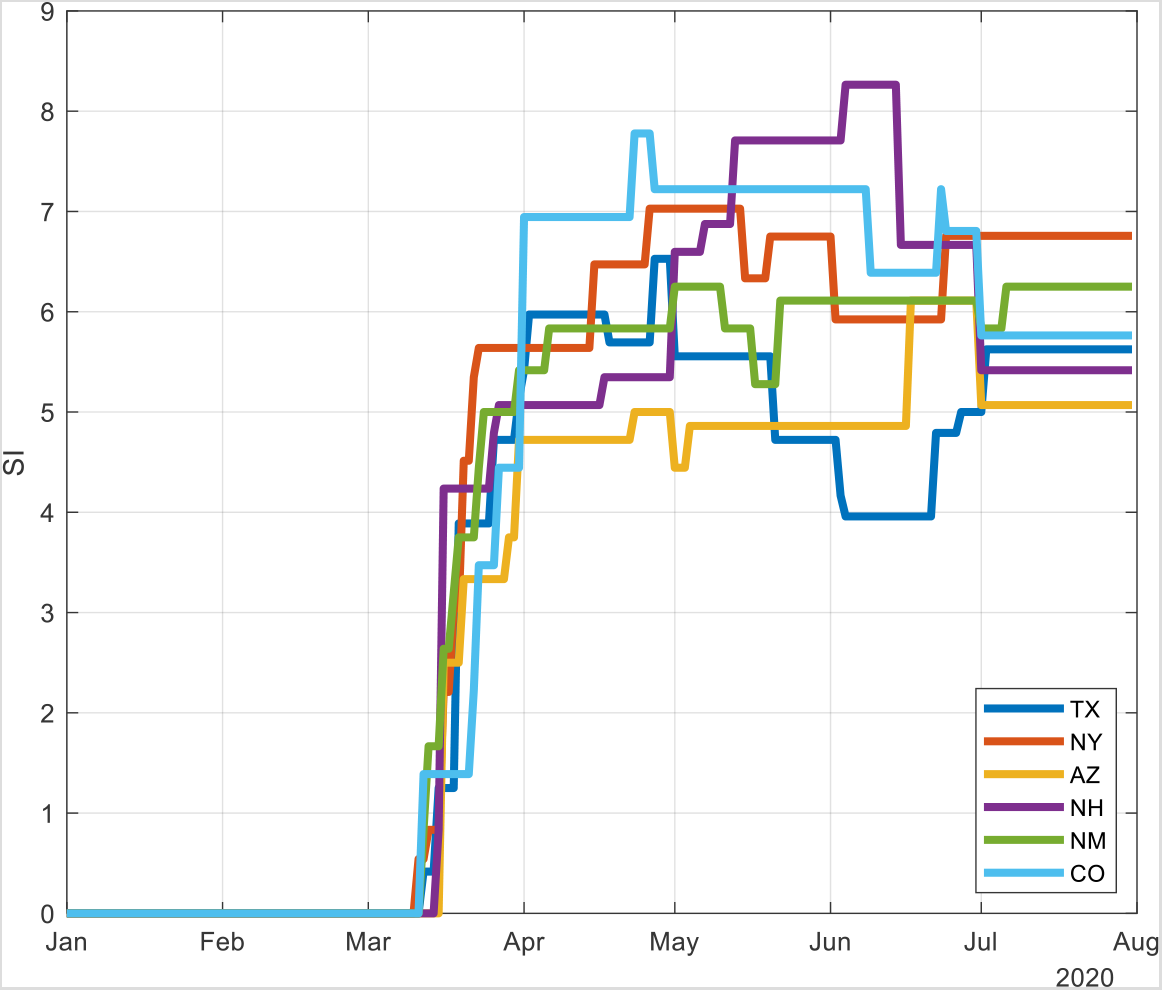
Path of Mitigation Measures (Bentley Stringency Index) over Time in each State.

## THE MODEL

The model that we adopt is based on the SEIRD (Susceptible, Exposed, Infected, Recovered or Died) model that has been developed by Weitz and Dushoff (2015), Loli and Zama (2020) and Lattanzio and Palumbo (2020). Diagrammatically it can be shown as follows:

**Figure 3.**
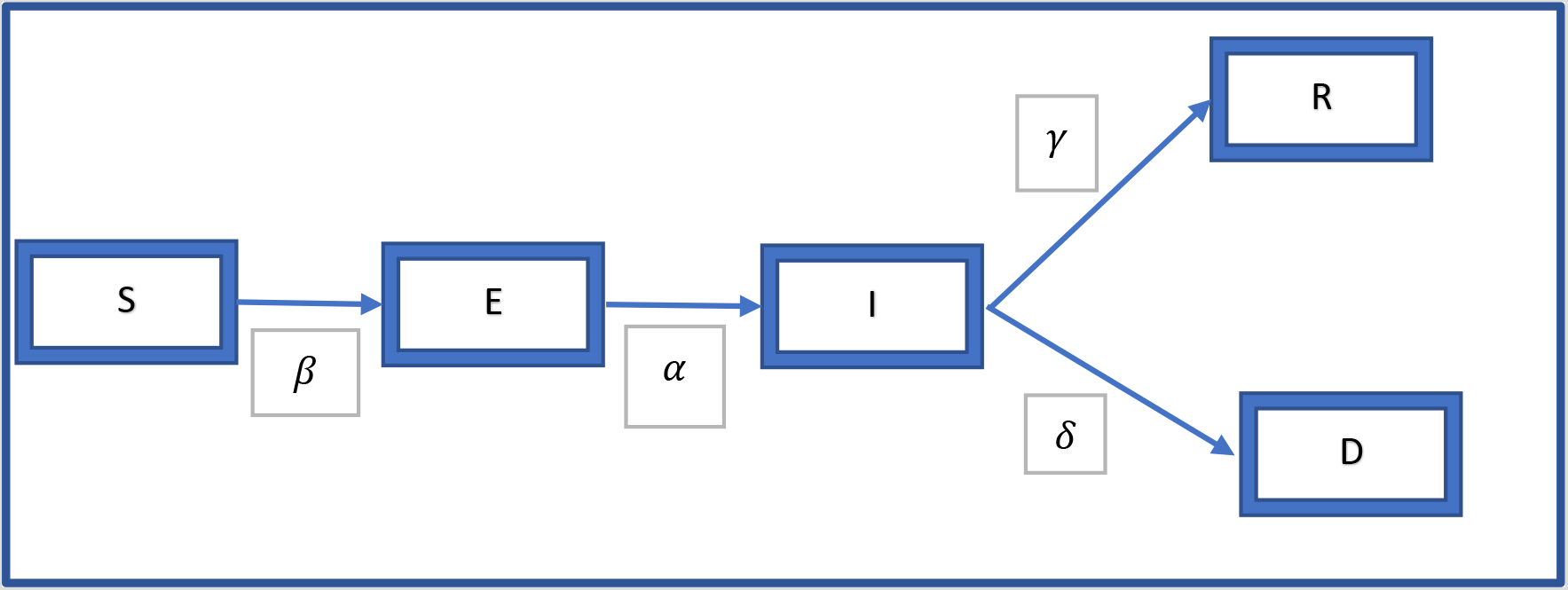
SEIRD MODEL FLOW CHART.

The structure of this model that is used in the literature can be described by the following equations:

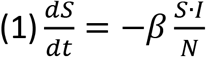

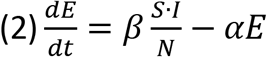

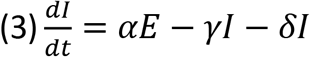

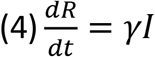

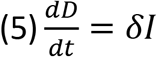

Following Lolli and Zama (2020), we have the compartments S, E, I, R and D. S is the susceptible group, E consists of those who are infected but may not be infectious, I contains the infectious group, while R and D respectively consist of those who recovered or died.

*β*: The term *β*, though called by slightly different names in the literature is often called the transmission rate of infection or the rate at which two specific individuals come into effective contact per unit of time^12^ (see Keeling and Rohani, 2011 and Vynnycky and White, 2010). Specifically, *β* is the product of the Contact Rate (average number of contacts per person per unit of time) * Transmission probability or probability of disease transmission in a contact.

*α*: The incubation rate, 1/*α* is the average period (days) of moving from E to I.

*γ*: The recovery rate for those infected and who have recovered. They move from I to R.

*δ*: The death rate of infected patients who die. They move from I to D.

1/(*γ* + *δ*): The average infectious period (days) for the infectious group I.

This model assumes that there is no reinfection and so eliminates movement from R to S. In addition due to the short period under consideration for epidemics, the population is assumed to be constant with equal birth and death rates.

The SEIRD model has now been modified by explicitly modeling two important drivers of *β*. In the epidemiological literature, it is acknowledged that *β* can depend on factors like age, living conditions, behavioral interventions such as the closing of theaters, schools, staggering of office hours as happened during the 1918 pandemic (Vynnycky and White, 2010; Bootsma and Ferguson, 2007; Hatchett et. al., 2007). Thus the standard *β* in the literature is a kind of “black box” which contains a complex of factors that can affect *β* and the transmission of disease. We investigate two important factors that influence *β*: mitigation policies that the BSI captures, and the compliance to these by the public. The latter is being increasingly discussed in the media as being crucial to the success of the various mitigation measures. On Aug 5, in a virtual symposium hosted by the Harvard University’s T.H. Chan School of Public Health, Dr. Fauci, Director of the National Institute of Allergy and Infectious Diseases, explained that it was due to the difference in the states’ mitigation measures and the different ways in which the public has complied with these measures that the U.S. is having difficulty in controlling the pandemic^13^.

To operationalize these two factors that drive *β* we now explicitly formulate a Mitigation Function that will act to reduce the transmission of disease. This *Mitigation Function* will have two major components^14^: the *Bentley Stringency Index* (BSI) and what we call the *Compliance Indicator* (CI). The BSI has been calculated daily for each state while the Compliance Indicator will be estimated from our adaptive computing procedure.

By explicitly including the Mitigation Function, we propose the following formula for *β*:

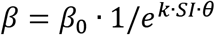

Where, *β*_0_ connotes the transmission without policy intervention, and *e*^*k·SI·θ*^ is defined as the mitigation function where k is 1/(average household size) and is fixed in our model estimation for each state, *SI* is the policy stringency index, and θ is the compliance indicator. *β* will decrease at an exponential rate of 1/*e*^*k·SI·θ*^.

To incorporate the transmission factor in a time-varying model, we develop a time dependent parameter *β*_*t*_ in our model for estimation purposes.

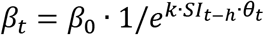

The time-varying Mitigation Function is: *e*^*k·SI*_*t−h*_·θ_*t*_^ where, *SI*_*t*_ is the stringency index at time t. The time lag introduced by a delay in policy implementation is denoted by *h*. We assume a modest policy lag of 1 day in our model using daily data. θ_*t*_ is the Compliance Indicator at time t.

Note that the Compliance Indicator can vary from 0 (no one complies) to a theoretical maximum of 1 (everyone complies). When the CI = 0, the BSI index has no effect irrespective of its value and the model collapses to the standard model where the mitigation efforts do not affect *β*. On the other hand, the BSI can also vary from 0 (where there are no restrictions) to a theoretical maximum of 10 (which is akin to a total lockdown). When BSI = 0, the model again collapses to the standard model. When BSI is greater than 0, then the effect on *β* will depend also on the Compliance Indicator. Even if BSI is at its maximum, a low CI will reduce the Mitigation Function. In other words, it is both BSI and the Compliance Indicator that will determine (given the household size that varies by state) the power of the Mitigation Function which affects the transmission of the disease. Incorporating this, our model structure will be as follows.

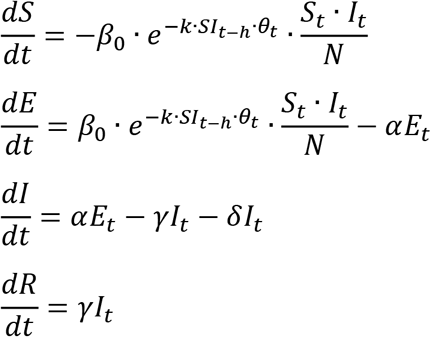

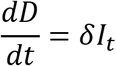

## METHODOLOGY AND RESULTS

To estimate the parameters of our proposed model, we use numerical analysis methods and statistical approaches with COVID-19 data from 6 states beginning from Mar 2020 to July 2020. In this computing process, we first develop the difference equation system as per the following system:

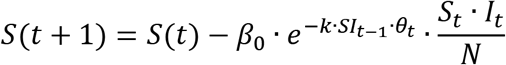

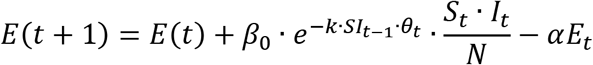

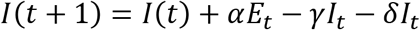

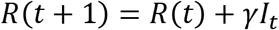

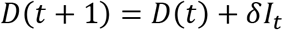

Then, we develop the overall error function: *Error*(*t*) = max{*Er*_*S*_(*t*), *Er*_*I*_(*t*), *Er*_*R*_(*t*), *Er*_*D*_(*t*)}, where *Er*_*S*_(*t*) = |*Ŝ*(*t*) − *S*(*t*)|; *Er*_*I*_(*t*) = |*Î*(*t*) − *I*(*t*)|; 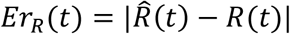; and finally, 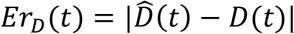.

As is commonly used, we take the absolute difference between *Ŝ*(*t*) and *S*(*t*) to denote the error *Er*_*S*_(*t*) at time t. We use a similar process to define the errors for Infection (*Er*_*I*_(*t*)), recovery (*Er*_*R*_(*t*)) and death (*Er*_*D*_(*t*)) at time t.

*Ŝ*(*t*), *Î*(*t*), 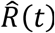, 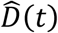 are the model estimations at time t for susceptible cases, confirmed infections, recovered individuals and those who died. Correspondingly, *S*(*t*), *I*(*t*), *R*(*t*), *D*(*t*) are the observed data for each compartment at time t. To find the estimated value**s** of the parameters at time t which give the minimum of *Error*(*t*), we applied the interior-point algorithms with dynamically modified constraints on the parameter estimations.

MATLAB 2020a was used for the computation. With the numerical estimation of the parameters, the 4^th^ order Rouge-Kouta method was used for predictions on *Ŝ*, *Î*, 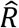, 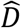 in all six states.

The model provides insights into the relationship between three crucial factors in controlling an epidemic: the Bentley Stringency Index (embodying the policy measures recommended or required by Federal or State authorities), the Compliance Indicator (embodying the extent of compliance by the public to these policy measures) and *R*_0_^15^ (the basic reproduction number).

With the estimated parameters, our next step explores the interaction of the time-varying values of the Stringency Index as well as the Compliance Index in each state with the movement of the infection rates.

To examine this, we did two sets of simulations to explore interacting effect of BSI and compliance rate on *R*_0_. Both simulations are based on the formula above, with fixed parameters of average household size (2.6 US average in 2019), *γ* and *δ* (from US data). *β*_0_ in this simulation is estimated from CDC’s report as of Aug 13^th^ (CDC, 2020).

### Simulation 1. Comparing the Oxford and Bentley Stringency Indices

Using our model, we calculated the movement of *R*_0_ using both the Bentley and the Oxford indices. In our model, the basic reproduction number

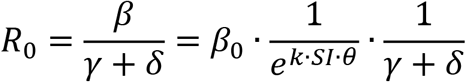

For an epidemic to die out, *R*_0_ must be less than one.

We ran the comparative simulations with fixed parameters, including the Compliance Indicator on all 6 states. The results (Figure 4) are reported for New York and Texas^16^. In this simulation, we assumed the CI to be a fixed value 0.5 for both states (Corona Board, 2020); average house hold size as 2.6; *γ* as 0.52 and *δ* as 0.03. Comparing the two Stringency Indices, the Bentley Stringency Index is overwhelmingly (with a few exceptions) more conservative in that the simulated *R*_0_ is higher than that obtained by using the Oxford Index.

**FIGURE 4:**
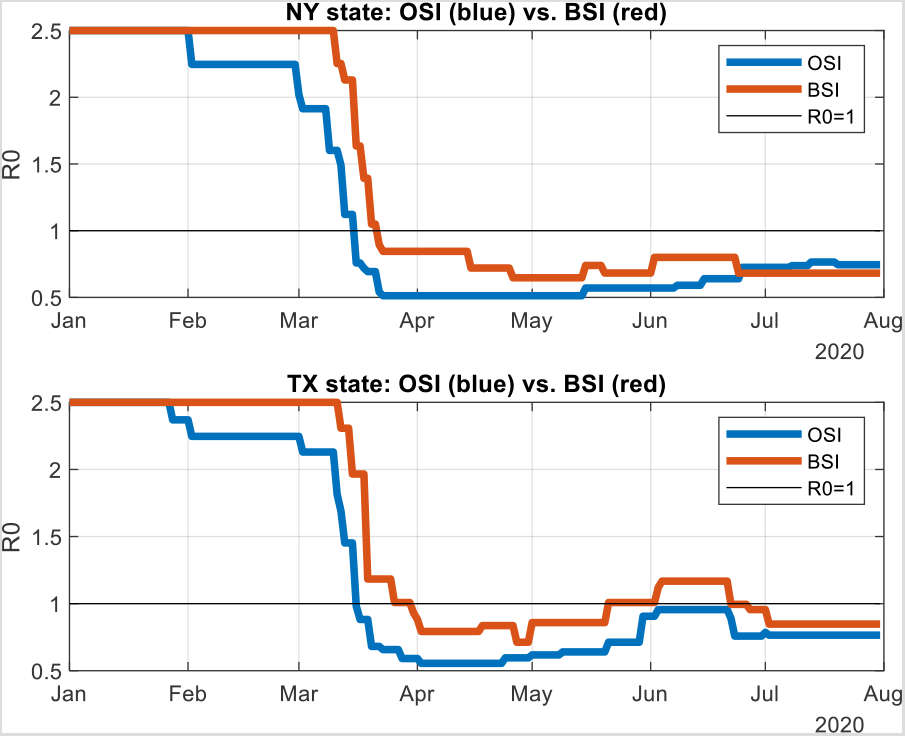
Comparison of Oxford Stringency Index (OSI) and Bentley Stringency Index (BSI)

These simulations gave us further confidence about using the Bentley Stringency Index. Recall that the BSI was constructed specifically for each state using the various components as described. In taking individual states separately, we were able to start with the first statewide announcement regarding mitigation measures (State of Texas, 2020) (State of New York, 2020).

### Simulation 2: Interaction of Compliance Indicator and *R*_0_ at different levels of BSI

Based on 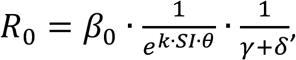 we simulated the Compliance Indicator and *R*_0_ at different levels of the BSI, assuming a recovery rate *γ* of 0.52 based on the U.S. average and an average household size of 2.6 (United Nations, 2019; Corona Board, 2020). See Figure 5.

**Figure 5:**
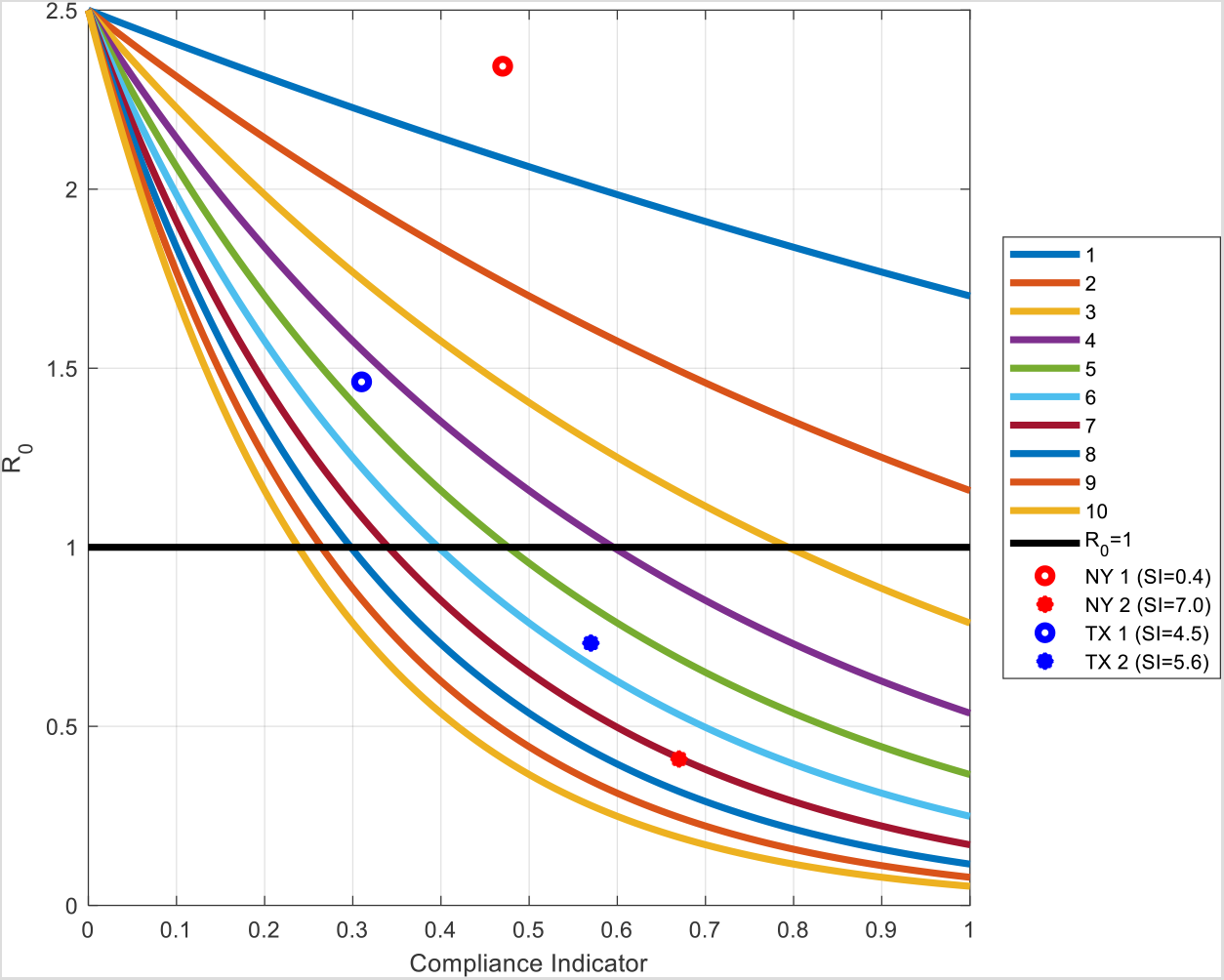
The Combination of Stringency and Compliance needed at various levels of R_0_.

In Figure 5, the vertical axis is *R*_0_ while the horizontal axis shows the range of the Compliance Indicator. Each colored line shows the relationship between *R*_0_ and the Compliance Indicator at different levels of the Stringency Index (The highest level of Stringency is 10 and is shown by the lowest line). The horizontal line denotes *R*_0_ = 1 which is accepted in the literature as the threshold above which the epidemic will keep spreading. When *R*_0_ is less than one (the lower part of the graph can be denoted as “desirable” and the upper portion as “undesirable”) then the epidemic will die out.

Our simulations show the different levels of compliance that would be compatible with different degrees of stringency in order for *R*_0_ to go below the threshold. For instance, when the BSI is at 1, then notice that even if the Compliance Indicator is at the maximum, *R*_0_ will not be lower than one. On the other hand, with the highest value of the BSI, the Compliance Indicator has to be at least 25% for *R*_0_ to be at the threshold.

Since the above figure is based on the average household size in the U.S., it must be remembered that when the household number increases, then to reach the same *R*_0_, the combination of the Stringency Index and the Compliance Indicator has to be at higher levels.

New York state began in the “undesirable” portion as the higher red dot indicates. That was during Day 65 (Mar 5th) to Day 75 (Mar 15th). However, between Day 120 (April 29^th^) to Day 130 (May 9^th^), New York moved to the “desirable” portion of the graph as the lower red dot shows. Similarly, Texas (blue dot) also moved from an undesirable point between Day 170 (June 18^th^) to Day 180 (June 28^th^) to the lower portion during the period Day 190 (July 8^th^) to Day 200 (July 18). We will be taking up these periods in more detail below.

This figure can be of use to guide policy regarding the extent of mitigation measures and compliance needed by the public. Let us illustrate what we mean by going back to the two states we showed in Figure 5, namely New York and Texas. These two states have had very different success in combating COVID-19. In Figures 6a and 6b we show plots of the estimated mitigation function against daily infections.

**Figure 6a New York:**
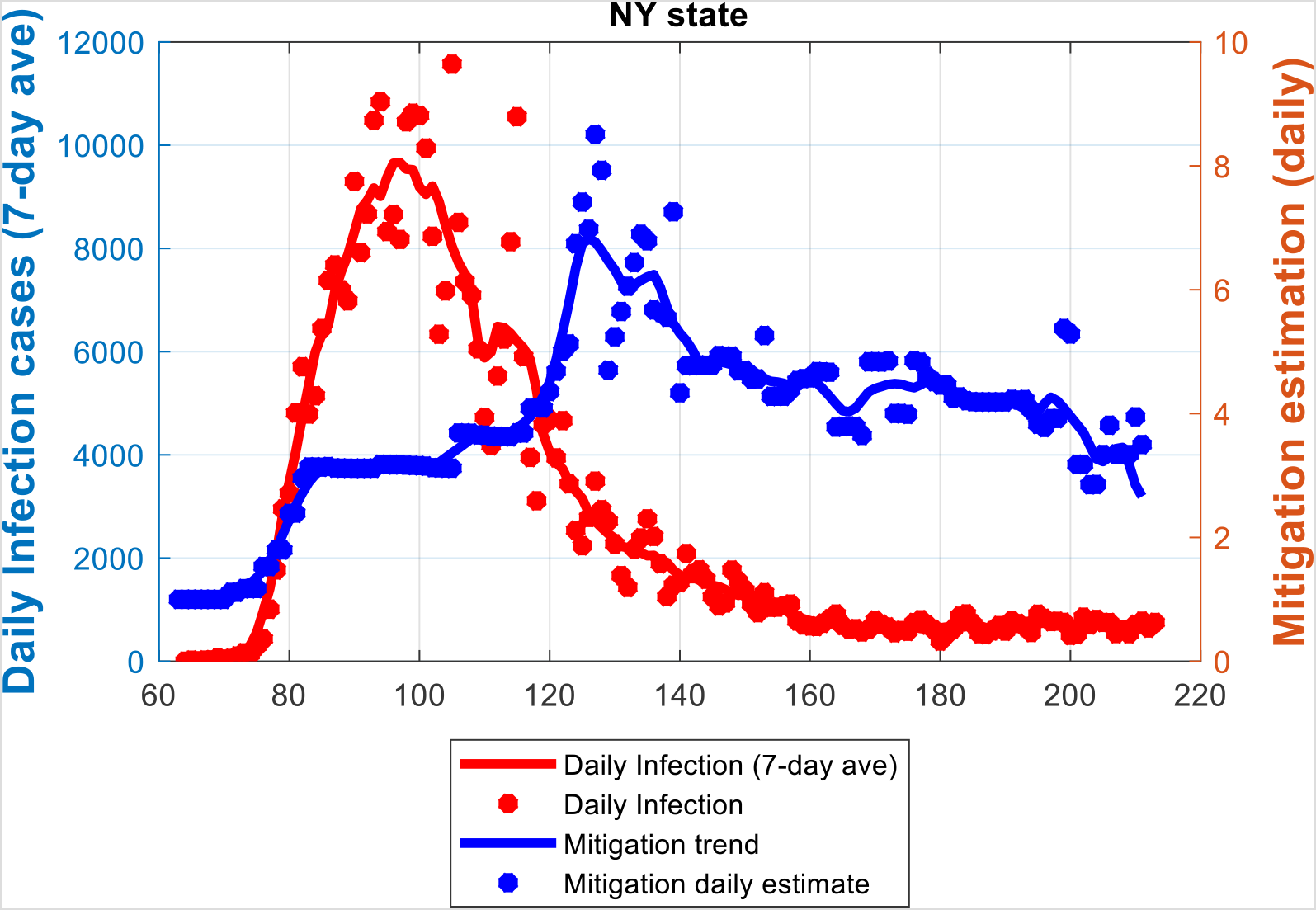
Plots of mitigation function estimation (daily) vs daily infections (7-day average)

**Figure 6b Texas:**
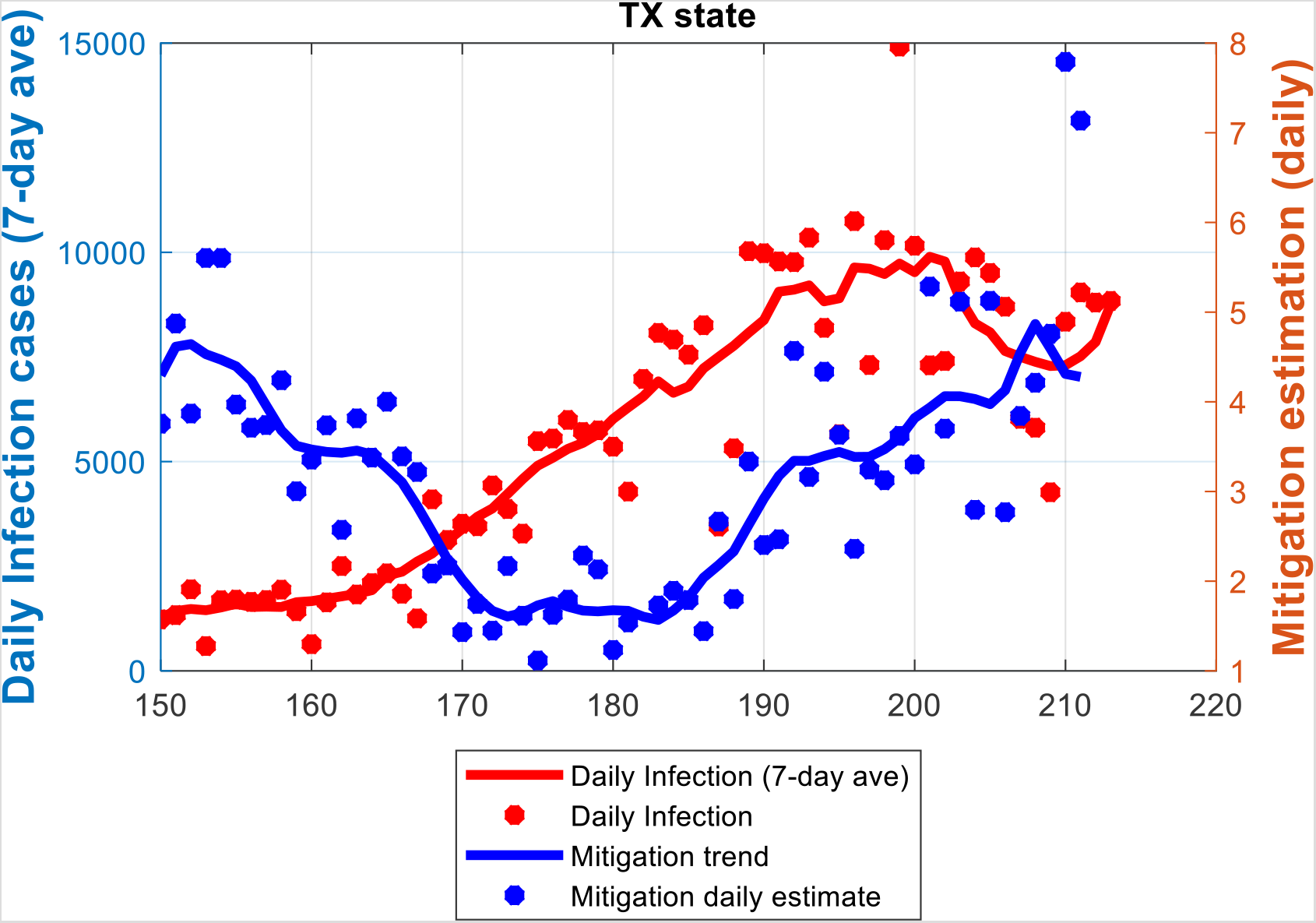
Plots of mitigation function estimation (daily) vs daily infections (7-day average)

In Figure 6a for New York, the inverse relation between the Mitigation Function (blue line) and daily infections (red line) can be clearly seen. It must be remembered that the Mitigation Function only captures a portion of all the factors that drive *β*, the transmission rate. When the infection was raging, the Mitigation Function was increasing and we find that the BSI increased from, essentially, 0 to 0.8 with a mean value of 0.358 during the 10-day period Mar 5^th^ to Mar 15^th^(Days 65 to 75). This was because New York recommended against nursing home visits, restricted public gatherings and cancelled public events. The mean estimated CI was 46%. This placed New York in the upper “undesirable” portion of the graph in Figure 5. When the state initiated the face mask requirement around April 15^th^ (Day 106) and the testing policy was introduced and defined on April 26 Day 117), infections which had hit a peak began a trajectory downwards until it plateaued around mid-June. By the beginning of May the infection had abated and the mean values of BSI and CI were 7.028 and 67% respectively during the 10-day period between April 29^th^ and May 9^th^. This trend corresponded with the announcement of New York’s COVID-19 Testing Policy. With this combination of BSI and Compliance Index, New York moved to the lower “desirable” range (Figure 5) as the infection rate came under control.

For Texas, on June 3^rd^ (day 155 in the figure) Governor Abbott signed an executive order on Phase III reopening, to allow more businesses to reopen and to reduce restrictions on gatherings. The Mitigation function decreased accordingly. Note in Figure 6b that as the Mitigation Function decreased with the relaxation of state restrictions, Covid-19 infections also began to climb.

Further, between Day 170 (June 18^th^) to Day 180 (June 28^th^), the BSI increased from 3.9 to 5.0 with a mean value of 4.5. The mean value of the estimated CI is 31%. Even though the state had increased the mitigation policies, it was not enough. The fact that the compliance also is far too low can be seen with a little thought experiment using Figure 5. Suppose the BSI were 5 (more stringent), then the compliance indicator would need to be at least 50% for the infection to die out as Texas moves to the desirable upper portion of the graph. On the other hand, with this low level of compliance, it can be confidently predicted that the infection will rise. In fact, in the following weeks between June 22 and July 17 the confirmed COVID-19 cases reached 14,916, the highest one-day mark for Texas.

Our results also indicate that when stringency measures are constant, changes in the Compliance Indicator is associated with changes in the infection rates. In Texas, from June 4^th^ to June 21^st^ (Day 156 to 173 since Jan 1^st^), the BSI stayed constant at 3.96. On the other hand, the estimated CI showed a dramatic 5-day average drop from approximately 88% to 39%. On June 4, Texas reduced restrictions on gatherings by allowing for indoor assemblies such as at places of worship, among local government operations, child care services and recreational sports for youths and adults. The daily infections during the corresponding period also jumped from 1649 to 4430. This is clearly visible in Figure 6b.

## DISCUSSION AND CONCLUSION

The objective of this paper was to examine how mitigation policies and compliance to these polices can combine to advance or frustrate the fight against the COVID-19 pandemic. Our concern arose due to states with similar community mitigation measures having very divergent trends in infection rates. To the best of our knowledge, there is no epidemiological model that would help us understand this phenomenon. In this paper we propose a simple modification to bring both mitigation policies and compliance into a standard epidemiological model and, in exploring their interaction, understand what would be the minimum levels of each that would lead each state to a point where the epidemic would die out.

To accomplish our purpose, we use a SEIRD model incorporating both mitigation measures and public compliance that are present in the “black box” that is *β*. In doing this, we build upon the work of Kurcharski et. al. 2020 where they suggest that a combination of methods (testing, tracing, physical distancing, self-isolation and quarantine) may to needed to reduce effective transmission so that the epidemic is contained. We go further by taking a vector of mitigation policy measures and encapsulating them into a Bentley Stringency Index (BSI) similar to that built by Oxford. We use their methodology and thus build on and extend their work. Tailoring our state stringency indices by incorporating directives like wearing face masks, restrictions on nursing home visits that are appropriate to U.S. states, we contribute to the literature.

The best of mitigation measures need to be actually followed if they are to be successful. To capture public compliance, a state-specific Compliance Indicator was estimated from the model using the daily COVID-19 data from each state. In this context, Cano et.al 2020 have modelled scenarios with different levels of what they have termed social distancing. However this term is also used interchangeably with lockdown. They show that the less seriously the public takes the lockdown measures, the longer the epidemic will take to resolve and the number of deaths will increase. By quantifying public compliance through the Compliance Indicator, we build the Mitigation Function that encompasses both the BSI and the CI. We demonstrate the association of the movement of infections with movements in the Mitigation Function, ceteris-paribus. Thus, it is the combined effect of the mitigation measures and the compliance that is key. Without either, there can be no containing the epidemic.

Compliance by the public to mitigation measures is largely exogenous in democratic societies. However, by coordinated and effective public information campaigns, through example, by helping people understand that obeying these restrictions is crucial to reclaiming their lives, compliance may be enhanced. In this regard, Arriola and Grossman’s work, though in the context of Africa, may be interesting (Arriola & Grossman, 2020). They wanted to see how the social identity of individuals could affect their compliance with advice from public health officials. In the U.S., some states have adopted the “stick” approach; California is cutting off power to those who are defying restrictions (Treisman, 2020). However, if the public understands that it is in their own self-interest, the degree of compliance can be increased with their cooperation.

We have also contributed by suggesting a practical, real-time, visual policy tool that can be used flexibly not only to monitor the progress in controlling the disease but also to adjust policy in a dynamic fashion. It can be used to support decisions in adjusting mitigation policies by taking into consideration the level of public compliance as well. This tool is also simple enough to be used to educate the public on the importance of compliance.

The chief limitation of our analysis is a problem that all researchers on COVID-19 have to contend with at this time with an ongoing deadly and fast-moving pandemic. Even though we had only 4% of our observation points that were missing or questionable, there are concerns regarding the overall quality of the data (General Accounting Office, 2020).

Our work has opened up several avenues of future research. Our method of using the Mitigation Function in SEIRD can be applied to other epidemiological models as well. In addition, this methodology can also be translated to other countries, thereby providing another tool to the authorities in combating this epidemic. We have also made the first step in attempting to quantify the factors that go into “black box” of *β* and hope that our work will stimulate further exploration.

## Data Availability

All the data used in this paper are public data. All the data resources are listed in the reference list in this manuscript.

## Acknowledgements

We thank Jason Wells, Gaurav Shah, and Maria Skaletsky for their unstinting help with software support. We also thank our student assistants Amitabh Agrawal and Piotr Kolodziej for assistance with data collection.

## ENDNOTES

1 The importance of full compliance, though generally acknowledged, can be seen in the control of the Ebola outbreak (Do & Lee, 2016) where even a day’s delay in full compliance could double the number of infections.

2 The positivity rates as on August 16,2020 are: New York (0.83); New Hampshire (1.34), New Mexico (2.59), Colorado (3.83), Texas (15.32) and Arizona (10.79).

3 (Anderson, Heesterbeek, Klinkenberg, & Hollingsworth, 2020) discuss the importance of various country-wide mitigation measures. (Kucharski, et al., 2020) investigate the effect of isolation, contact tracing, testing and physical distancing on reducing transmission. See also (Hellewell, et al., 2020) and (Prem, et al., 2020). (Hatchett, Mecher, & Lipsitch, 2007) and (Bootsma & Ferguson, 2007) inform us on public health intervention measures during the 1918 pandemic to give us a historical context.

4 (Teslya, et al., 2020) showed the importance of mask wearing and hand washing in conjunction with social distancing. Mask wearing and handwashing are two measures entirely within the control of the individual, whereas other measures may need the cooperation of others. However, individuals may also decide not to comply with these measures and that is why our compliance index is so important.

5 See (Heesterbeek, 2002) and (Delamater, Street, Leslie, Yang, & Jacobsen, 2019) for a history of the basic reproductive number and its complexities.

6 As explained by (Chowell & Nishiura, 2008) a fundamental result in epidemiology is the “threshold” value of the basic reproduction number Ro: “ *There is a difference in epidemic behavior when the average number of secondary infections caused by an average infective during his/her period of infectiousness, called the* basic reproduction number*, is less than one and when this quantity exceeds one”*.

7 (State of New Hampshire, 2020) (State of Colorado, 2020) (State of New Mexico, 2020) (State of Texas, 2020) (State of Arizona, 2020) (State of New York, 2020)

8 (Nande, Adlam, Sheen, Levy, & Hill, 2020) found that within-household transmission was an important element for success in controlling the infection. In this context, see also (Li, et al., 2020). (Emeruwa, et al., 2020) in a study of nearly 400 pregnant women in New York City did not find an association between infection and population density but did find a higher risk of COVID-19 infection due to increased household crowding.

9 (Stutt, Retkute, Bradley, Gilligan, & Colvin, 2020) showed the effectiveness of wearing face masks in managing the Covid-19 pandemic.

10 (CDC, 2020) Some are advocating the term “physical distancing” to clarify the difference between physical and social distancing where social connectivity is to be encouraged while yet maintaining physical distancing. See (Allen, Ling, & Burton, 2020)

11 As per CDC guidelines (CDC, 2020)

12 It is important to clarify that we are talking of a frequency-dependent transmission in that the number of contacts does not depend on population size. See (Keeling & Rohani, 2011).

13 In the words of the Harvard Gazette, “In addition, he (Dr. Fauci) said, state reopening plans proceeded at different paces. Some states reopened slowly, similar to the pace of European nations, while others went much faster. Another variable, he said, was the extent to which residents of different states adhered to reopening guidelines, with some following recommendations while others ignored the restrictions, sometimes in notably large groups”. (Powell, 2020)

14 The third component is household size that we will be taking as constant over the period under consideration.

15 Ro is the basic reproductive rate and is defined in the literature as the average number of people an infectious person will infect assuming that the rest of the population is susceptible.

16 This is in the interest of space. Results for other states are available on request.

17 Targeted refers to specific areas of the state the policy applies, while general indicates the policy is implemented statewide.

## APPENDIX A

**Table XX.**
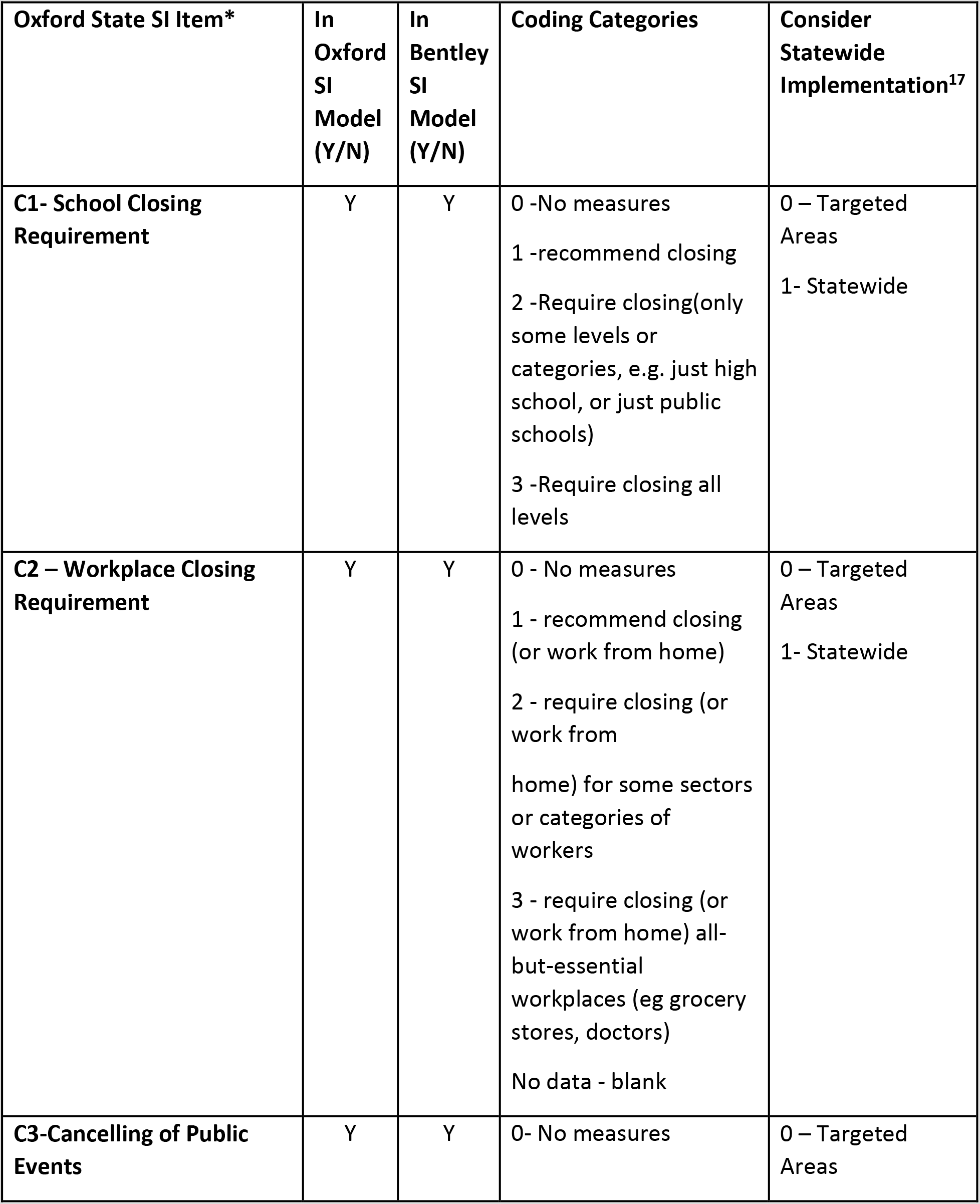
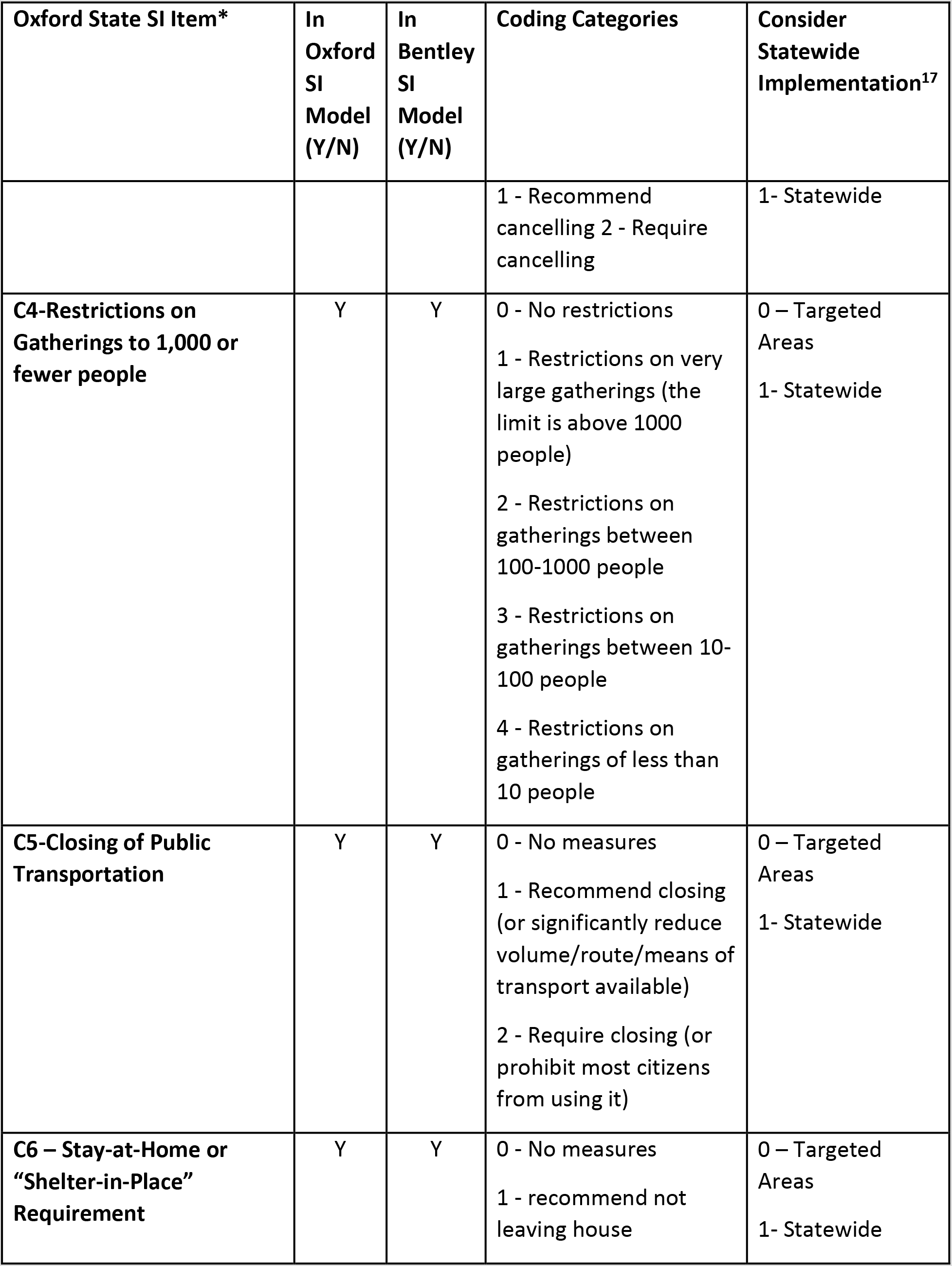
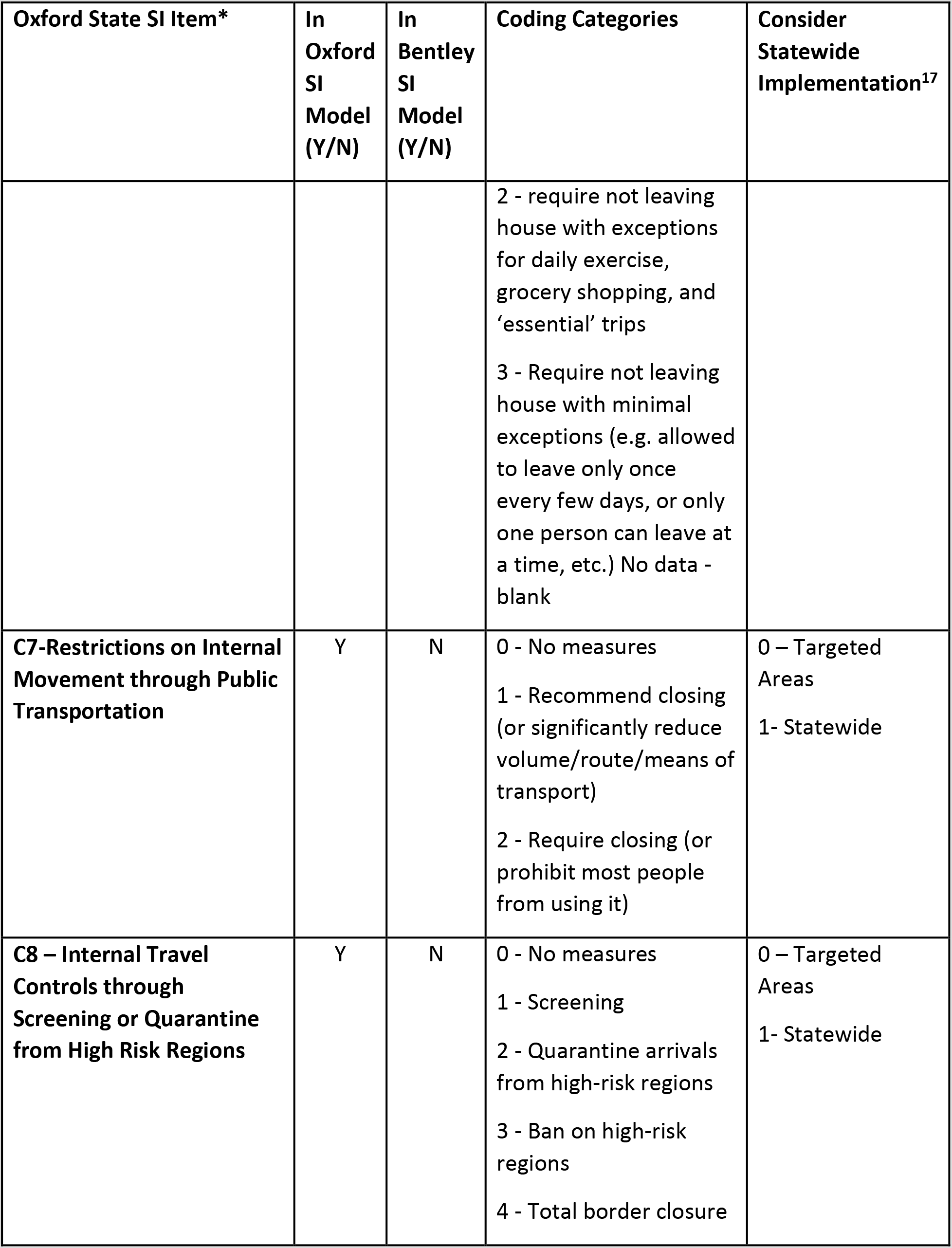
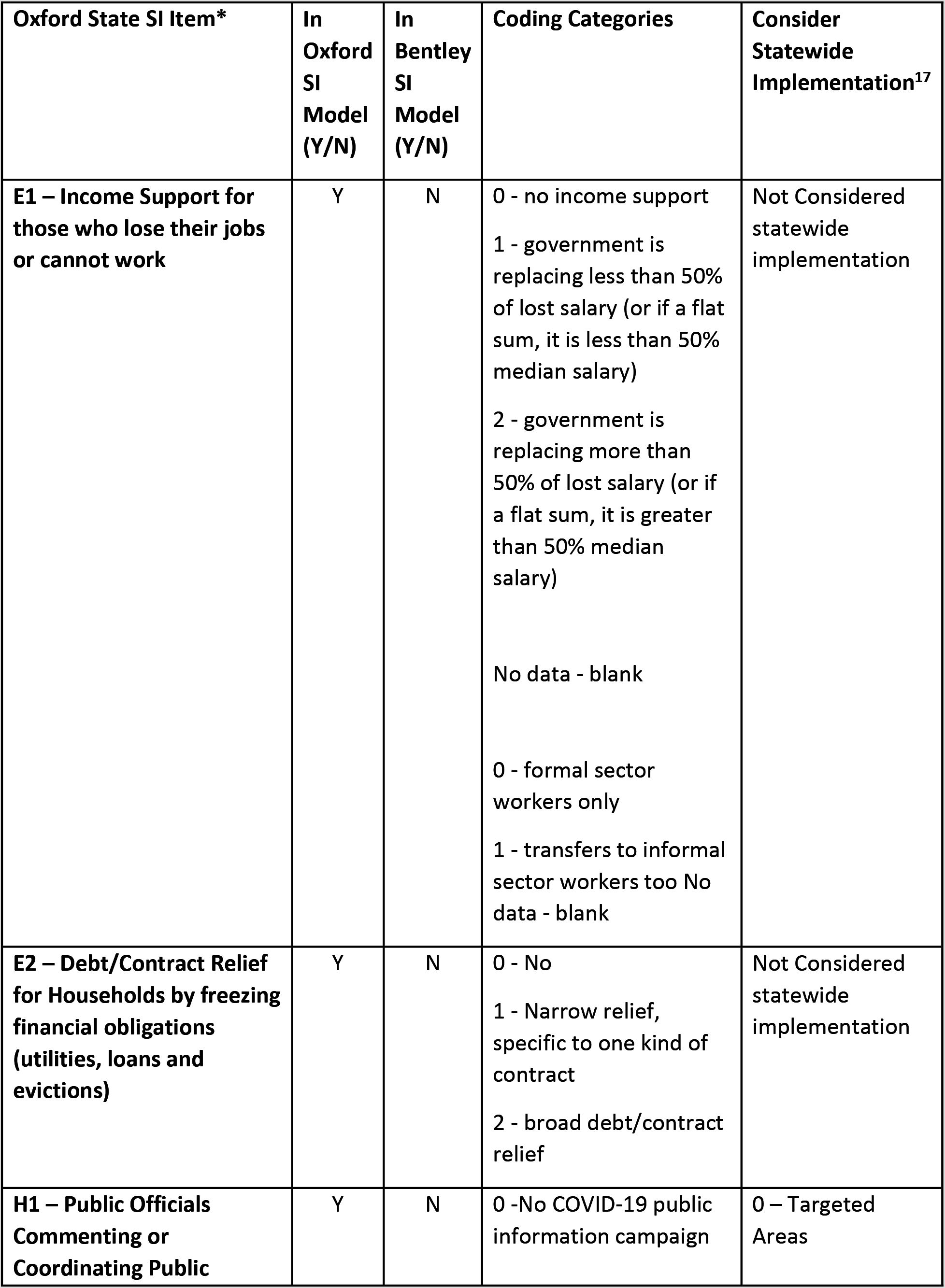
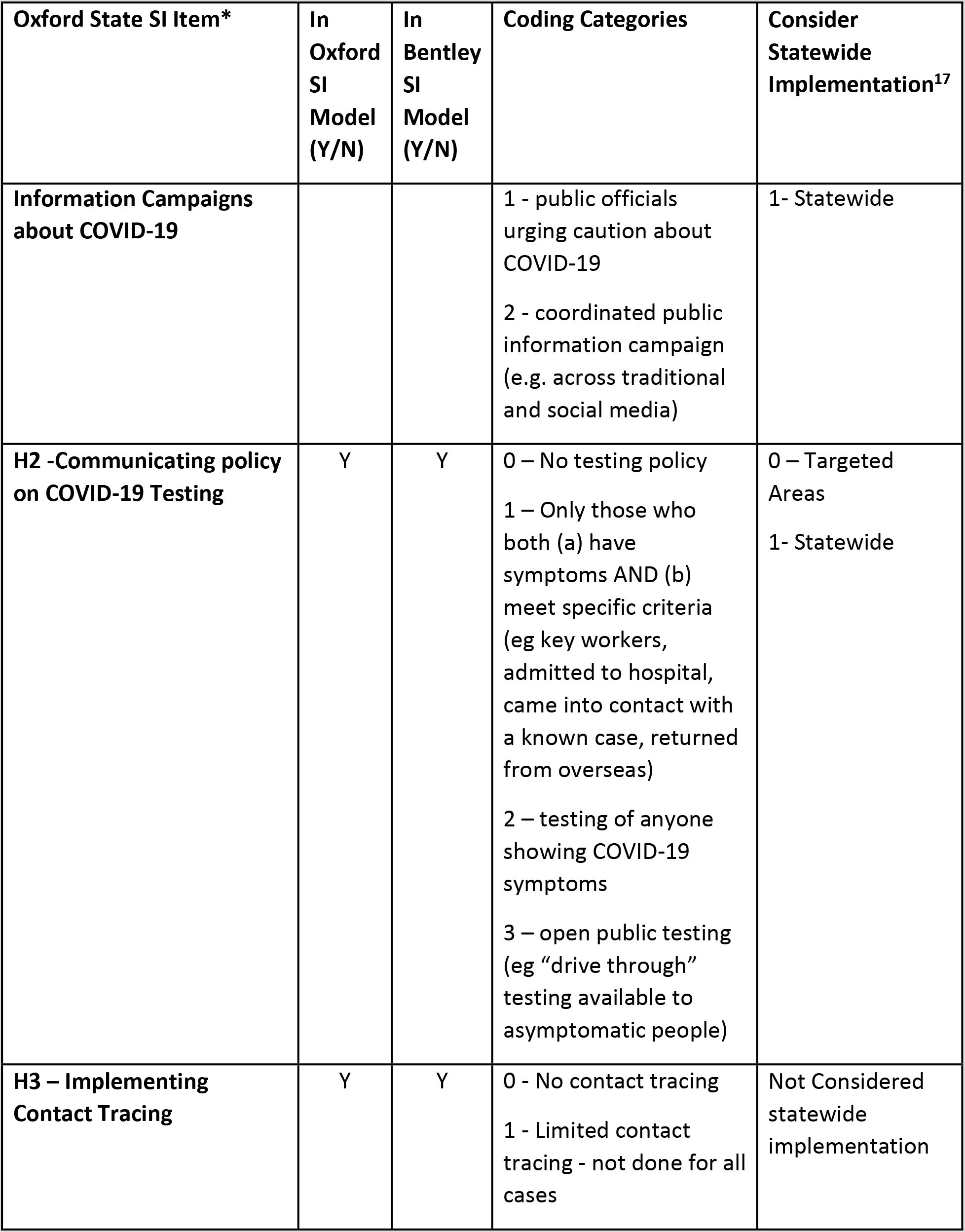
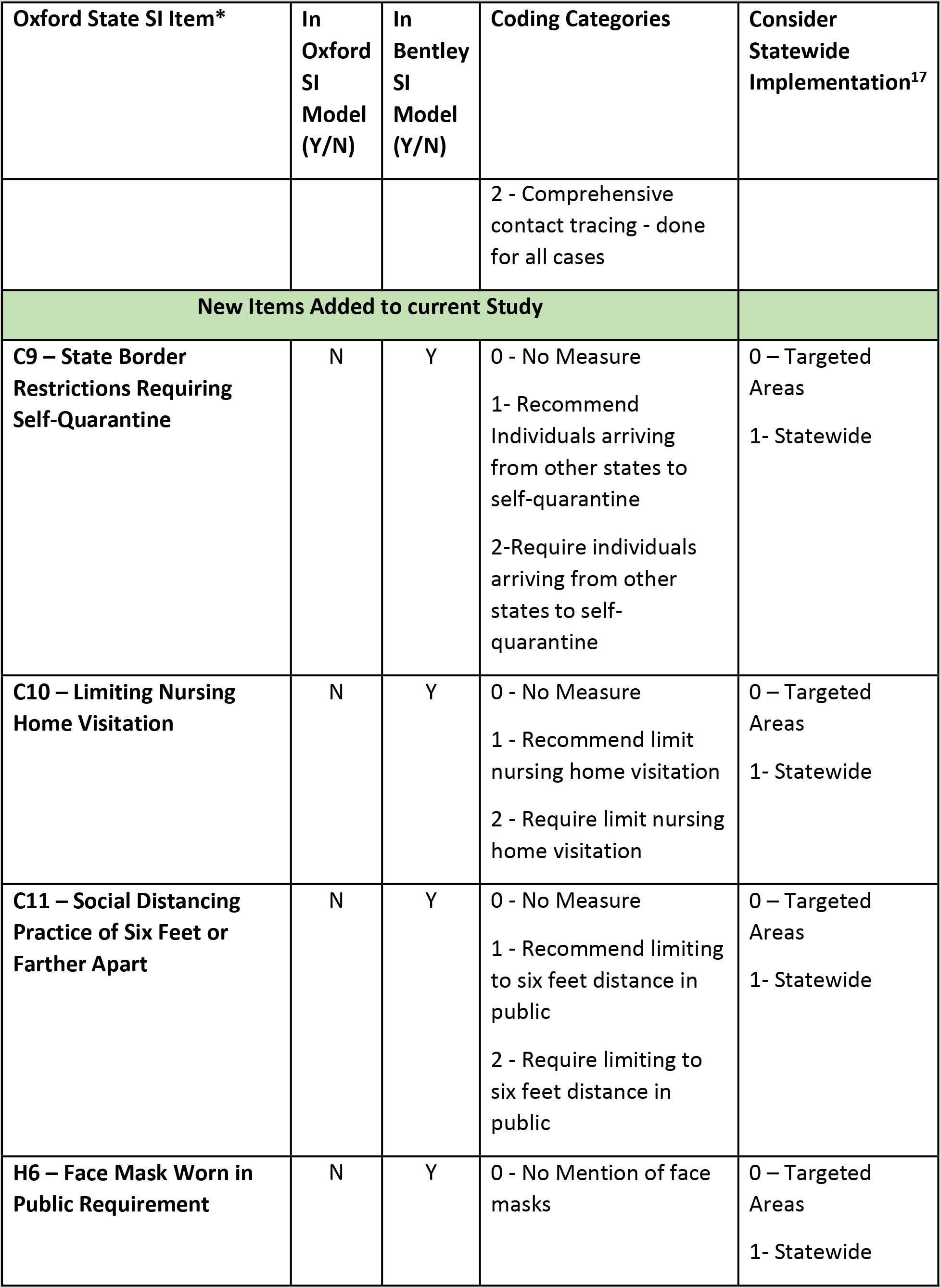
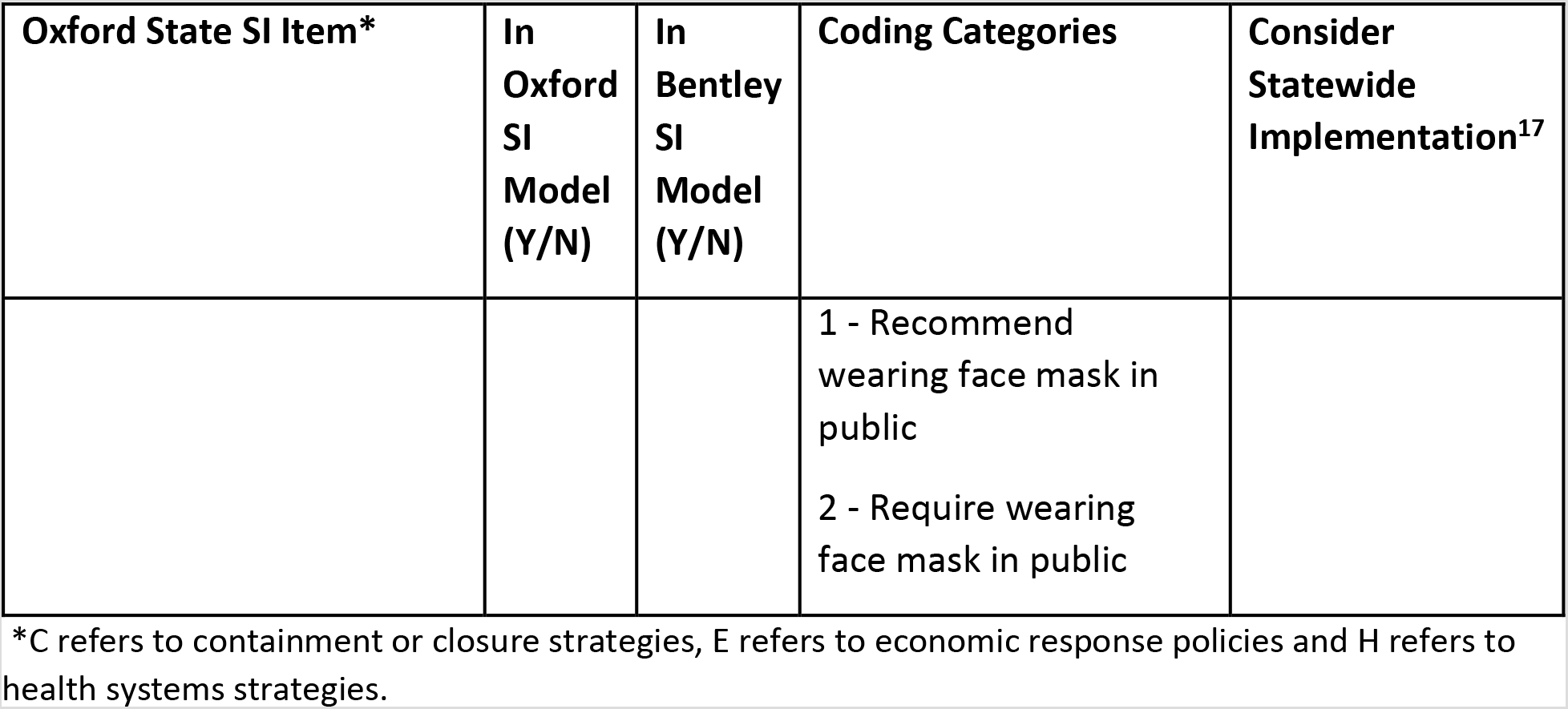
Items included in the Oxford and Bentley state specific Stringency Indices.

## Appendix B Creation of new items in the Bentley modification of the Oxford Stringency Index.

Our state stringency index (SI) is compiled from 12 items. The Oxford state SI is based on 13 items (Hale, et al., 2020; Hale, et al., 2020B). We modified the Oxford state stringency index by dropping five items and adding four. We dropped item C7 (restriction on internal movement through public transportation) because this policy was already included in item C5 (closing of public transportation). We also dropped C8 (internal travel controls through screening or quarantine from high risk regions) because we were not aware of states closing borders entirely or banning the entry of individuals from high risk regions. Even Rhode Island’s attempt to target cars with New York state license plates in order to impose a 14 day quarantine on visitors from New York was rescinded (Moroney, 2020). We also set aside items E1 (income support for those who lose their jobs), E2 (debt/contract relief for households) and H1 (public official commenting or coordinating public information campaigns about Covid-19) because we felt they did not pertain to containment or closure. We added a total of four items to the stringency index. These items pertain to states imposing a mandatory self-quarantine on visitors from other states (C9), limiting nursing home visitation (C10), recommending a social distancing practice in public of at least six feet (C11) and policy on face mask covers (H6).

The daily stringency index score is tabulated as an average of 12 sub-indices drawn from the containment and closure and health systems items in the Oxford inventory scaled from 0 to 100. Additional weight is given to 10 of the 12 policy items that could potentially be implemented statewide. Because the four items we created (see Appendix A Table) resulted in two more items than the original Oxford SI that could be implemented statewide, we modified the weight on which the final stringency index score is based. This is done by weighing the Likert scale points of the 10 indicators that are designated for statewide or targeted implementation using the formula outlined by the Oxford group.

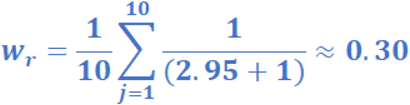

Our weight of 0.30 was very close to the ones calculated by Oxford (0.29), based on eight items, for use in the original index. The weights are then employed to create sub-indices of those items scaled to 100 in the same manner indicated from Oxford.

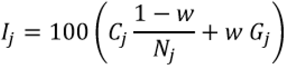

For the two items without statewide implementation as an option, sub-indices were calculated simply as a scaled function of 100 multiplied by the ratio of the raw scores to their maximum point value.

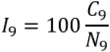

The final Stringency Index score is then an average of the all sub-indices.

## Notes

### Competing Interest Statement

The authors have declared no competing interest.

### Funding Statement

This paper is not supported by any funding.

### Author Declarations

This paper is based on public data. There is no human subjects or any clinical trials involved.

